# Cabergoline as a Preventive Migraine Treatment: An Investigator-Initiated Randomized Controlled Trial

**DOI:** 10.1101/2023.12.27.23300273

**Authors:** Astrid Johannesson Hjelholt, Flemming Winther Bach, Helge Kasch, Henrik Støvring, Troels Staehelin Jensen, Jens Otto Lunde Jørgensen

## Abstract

**Background:** Beneficial effects of dopamine agonist treatment on migraine have been reported but remain to be properly tested.

**Aim:** to examine the effect of cabergoline as preventive treatment for migraine.

**Primary endpoint:** Change in monthly migraine days (MMD).

**Methods:** In a randomized, double-blind, placebo-controlled pilot study, 36 adults with episodic and chronic migraine were enrolled. Following a 28-days baseline period, participants received cabergoline 0.5 mg or placebo once weekly for 12 weeks as add-on treatment. An electronic headache diary was completed by the participant, and pertinent headache questionnaires and blood tests were collected at baseline and following the treatment period. The trial was registered with ClinicalTrials.gov (NCT05525611).

**Findings:** Mean (SD) baseline MMD was 13.6 (4.1) in the cabergoline group and 14.0 (5.3) in the placebo group. In participants with episodic migraine (n= 20), the change in mean MMD (SE) from baseline to the last 28 days of the treatment period was -5.4 (1.3) (cabergoline) as compared to -1.8 (0.9) (placebo) [odds ratio: 0.79 (95% CI 0.65 - 0.95), p=0.014]. In participants with chronic migraine (n=13), the reduction in MMD with cabergoline was not significant (p=0.6). Patients’ global impression of change significantly improved after cabergoline as compared to placebo in the entire group of participants (p=0.006). The number of participants with episodic migraine achieving ≥ 50% reduction in MMD tended to increase after cabergoline (p=0.07). Seven participants receiving cabergoline and 4 participants receiving placebo experienced adverse effects, none of which were serious.

**Interpretations:** Preventive cabergoline treatment exhibited clinically meaningful improvement in episodic migraine without serious adverse effects. This provides proof-of-concept to justify a sufficiently powered phase 2 trial with different cabergoline dosing regimens as preventive treatment of episodic migraine.

**Funding:** This study has received no external funding.

## Introduction

Migraine is a complex neurovascular disorder characterized by recurrent attacks of headache, which are typically unilateral, pulsating and accompanied by nausea, vomiting, and photo- and phonophobia^1^. The estimated global lifetime prevalence is ≈ 15%, with a two- or threefold higher frequency in women. According to the Global Burden of Disease 2019 study, migraine is the second leading cause of disability worldwide, quantified as years lived with disability^2^. The mechanisms contributing to the pathophysiology and sexual dimorphism of migraine are not fully understood, but nociceptive signals from the trigeminovascular system in combination with vasoactive compounds such as calcitonin gene-related peptide (CGRP) are considered pivotal^1^.

The pharmacological treatment includes acute migraine–specific medication, in particular selective serotonin 5-HT1B and 5-HT1D receptor agonists, as well as preventive treatments aiming to reduce frequency, duration and severity of attacks^1^. Therapies blocking the CGRP pathway have proven effective as both acute and preventive treatment, but the cost is a constraint, and they are not effective in all patients^3^.

Ergot alkaloids, which were previously used as migraine treatment, are vasoconstrictors of the carotid artery beds and activate 5-HT as well as dopamine receptors^4^. The role of dopamine in migraine pathophysiology, however, remains ambiguous. Dopamine antagonistic drugs are used for acute treatment of migraine and certain premonitory symptoms of migraine are considered to be dopamine-driven^5^. On the other hand, the dopamine D2 receptor agonist bromocriptine, an ergot alkaloid derivative, and other dopamine receptor agonists have shown beneficial effects in migraine^6,7^.

A major indication for dopamine agonists, including bromocriptine, quinagolide and cabergoline, is hyperprolactinemia usually caused by a prolactin-secreting pituitary adenoma. Interestingly, hyperprolactinemia is more prevalent in females and often associates with headache, which is frequently alleviated by dopamine agonist treatment^8–11^. In a case series, we observed hyperprolactinemia-associated unilateral headache including migraine to resolve in a rapid and pronounced manner after dopamine-agonist treatment irrespective of adenoma size and prolactin lowering^8^. In line with this, preclinical and clinical studies indicate a link between prolactin and migraine^12^. Prolactin interacts with neural circuits including trigeminal sensory neurons, where it increases neuronal excitability^12^. In addition, moderately elevated prolactin levels are reported in migraine patients, in whom uncontrolled dopamine agonist-induced prolactin lowering improves headache in a high proportion of patients^13,14^. Cabergoline, which is also a ligand for the 5-HT1B and 5-HT1D receptors, has fewer side effects compared to other dopamine agonists and its pharmacokinetic properties allow weekly oral administration^15,16^.

The aim of this investigator-initiated, randomized, placebo-controlled pilot study was to examine the effect of cabergoline as preventive treatment for migraine. The primary endpoint was change in monthly migraine days (MMD).

## Method

### Trial design

This pilot study was an investigator-initiated, randomized, double-blind, placebo-controlled, parallel trial conducted at Aarhus University Hospital from November 2022 to June 2023. It comprised two phases: a 28-days baseline period followed by a 12-week treatment period. Prior to the 28-days baseline phase, participants were randomly assigned in a 1:1 ratio to receive either cabergoline 0.5 mg or placebo once weekly as an add-on treatment. The pharmacy at Aarhus University Hospital undertook randomization, blinding and provision of cabergoline and placebo. Block randomization was used, and the allocation sequence was stored in sealed envelopes until the study was unblinded after last participant’s last visit. Study data were collected and managed using REDCap (Research Electronic Data Capture) hosted at Aarhus University ^17^.

The study protocol was approved by the regional Ethics Committee System and the Danish Medicines Agency and reported at http://www.clinicaltrials.gov (NCT05525611). The study was conducted in accordance with Good Clinical Practice (GCP) under the supervision by the regional GCP unit. Written and oral consent was obtained from all participants.

### Trial population

Thirty-six adults (> 18 years of age) with a history of episodic migraine (< 15 MMD) or chronic migraine (≥ 15 MMD) were enrolled. Migraine was defined in accordance with the International Classification of Headache Disorders, 3rd edition^18^. A minimum of 6 MMD was required. The exclusion criteria included pregnancy, breastfeeding, use of drugs with dopamine antagonistic properties, heart valve disease, severe untreated hypertension and psychiatric disorder requiring treatment. In addition, fertile women without safe contraception (hormonal contraception and intrauterine devices) were excluded. Participants were not allowed to change their preventive migraine treatment during the trial, and patients with planned medication changes were not included. Also, patients with chronic daily headache including presumed medication overuse headache were not included.

### Trial assessments and safety evaluations

The following information was retrieved daily via an electronic diary during both the baseline period and the treatment period: migraine headaches including time of onset and resolution, pain severity (mild, moderate, severe) and features (unilateral or bilateral location, pulsating quality, aggravation by physical activity), associated symptoms (nausea, photo- and phonophobia, and symptoms of aura), use of acute migraine-specific and analgesic medications, missing workdays due to headache, contact to a general practitioner or the emergency department due to headache, and potential triggering factors.

At baseline and following the 12-week treatment period, the participants completed the Migraine Disability Assessment Score (MIDAS) and the Headache Impact Test-6 (HIT-6) questionnaires. MIDAS is developed to assess migraine-related disability over a 3-month recall period. It contains five questions regarding the number of days of missed work/school, reduced productivity at work/school, missed household work, reduced productivity in household work, and missed family and/or social activities. The total score is the sum of the five items with a higher score indicating greater disability^19^. The HIT-6 consists of six items: pain, social functioning, role functioning, vitality, cognitive functioning, and psychological distress. The answer to each of the six questions generates a total HIT-6 score from 36 to 78, where a higher score indicates a greater impact of headache on the daily life.

At the end of the treatment period, the participants reported their impression of change in overall disease status since the start of the study using the single-item patient global impression of change (PGIC) scale. The PGIC uses a 10-point scale ranging from “very much worse” (1) to “no change” (5), and “very much improved” (10). In addition, the participants were asked to provide their best guess as to whether they had received active or placebo treatment.

Blood samples were drawn at baseline and following the 12-week treatment phase and analyzed for prolactin. At baseline, fertile women underwent a qualitative human chorionic gonadotropin (hCG) blood test to rule out pregnancy.

Safety was monitored throughout the trial including reporting of adverse events and serious adverse events.

### Endpoints

The primary endpoint was change in MMD between the baseline period and the final 28 days of the treatment period. A migraine day was defined as any calendar day on which the participant had onset, continuation, or recurrence of migraine as recorded in the electronic diary. MMD was defined as number of migraine days per 28 days. Secondary endpoints included ≥ 50% reduction in MMD, PGIC, use of acute migraine–specific medication (triptans) and change from baseline in MIDAS score and HIT-6 score.

### Statistical analyses

Participants were analyzed as a whole group (entire) and stratified into participants with episodic migraine (6-14 MMD at baseline) and participants with chronic migraine (> 15 MMD at baseline), respectively. Change in MMD was calculated as the difference between number of migraine days per 28 days at baseline and the final 28 days of diary completion in the treatment period. In case of missing data due to lack of diary completion in the baseline period, a predicted value was imputed by dividing number of migraine days with number of days with diary completion multiplied by 28. The maximum number of missing diary completions for a participant in the baseline period was 4 days. The treatment effect was calculated by subtracting the MMD change in the placebo group from the change in the cabergoline group. The primary and secondary endpoints were analyzed using a linear regression model with heteroscedasticity-robust standard errors, computed via the HC1 method, or unpaired t-test and presented as mean ± SE if data were normally distributed. Non-normally distributed data were analyzed using Mann Whitney U test and presented as medians (IQR). To test for normality, Q-Q plots were evaluated in addition to the Shapiro-Wilk normality test. As a relative measure of effect, an unadjusted random effect logistic regression model was applied using follow up time as explanatory variable. This model accounts for differences in baseline proneness to attacks. The estimated odds ratio (OR) denotes the odds of a participant in the cabergoline group having a day with migraine, using the placebo group as reference. Binary variables were compared with Fisher’s exact test. Correlation analyses were performed using Spearman’s rank-order correlation. All statistical analyses were performed using R (version 4.3.0). Results were considered statistically significant at a significance level < 0.05 (two-tailed).

## Results

### Participants

Between September 13, 2022, and February 9, 2023, 101 adults were assessed for eligibility. Sixty-five were excluded, and 36 were randomly assigned to cabergoline or placebo (Supplemental Figure 1). All randomized participants completed the trial. In the final analyses, 3 participants were excluded (1 in the cabergoline group and 2 in the placebo group) due to too few MMD during the baseline period according to the inclusion criteria (Supplemental Figure 1).

Baseline characteristics of the participants are presented in Table 1. Twenty out of 33 participants had episodic migraine, and 13 had chronic migraine. The two groups were comparable except that the age at disease onset was lower in the cabergoline group among participants with chronic migraine. Thirty of the 33 participants were females. The mean age of the participants was 42.9 years with a mean age at migraine onset of 17.2 years. All participants used acute medications, and 30 used migraine–specific medications (triptans). Twenty-two participants were currently using or had previously used migraine-preventive medication; 19 had discontinued use of a preventive medication because of insufficient efficacy or side effects. During the baseline phase, the mean (SD) number of migraine days/28 days was 13.6 (4.1) in the cabergoline group and 14.0 (5.3) in the placebo group.

### Change in number of monthly migraine days

In the entire group of participants, no difference in MMD change between cabergoline and placebo was found [median (IQR): -5 (6) days (cabergoline) vs. -3.7 (4) days (placebo), p=0.4] (Table 2, Figure 1). In participants with episodic migraine, cabergoline significantly reduced the mean number of MMD from baseline as compared to placebo [mean (SE): -5.4 (1.3) (cabergoline) vs. -1.8 (0.9) (placebo); mean (95% CI) difference from placebo was -3.6 (-7.0, -0.2) (p = 0.04)] (Figure 1). In participants with chronic migraine, there was no significant change in number of MMD during cabergoline treatment as compared to placebo [median (IQR): -6.5 (7.5) days vs. -4.9 (7.5) days, (p=0.6)] (Figure 1).

**Figure 1.**
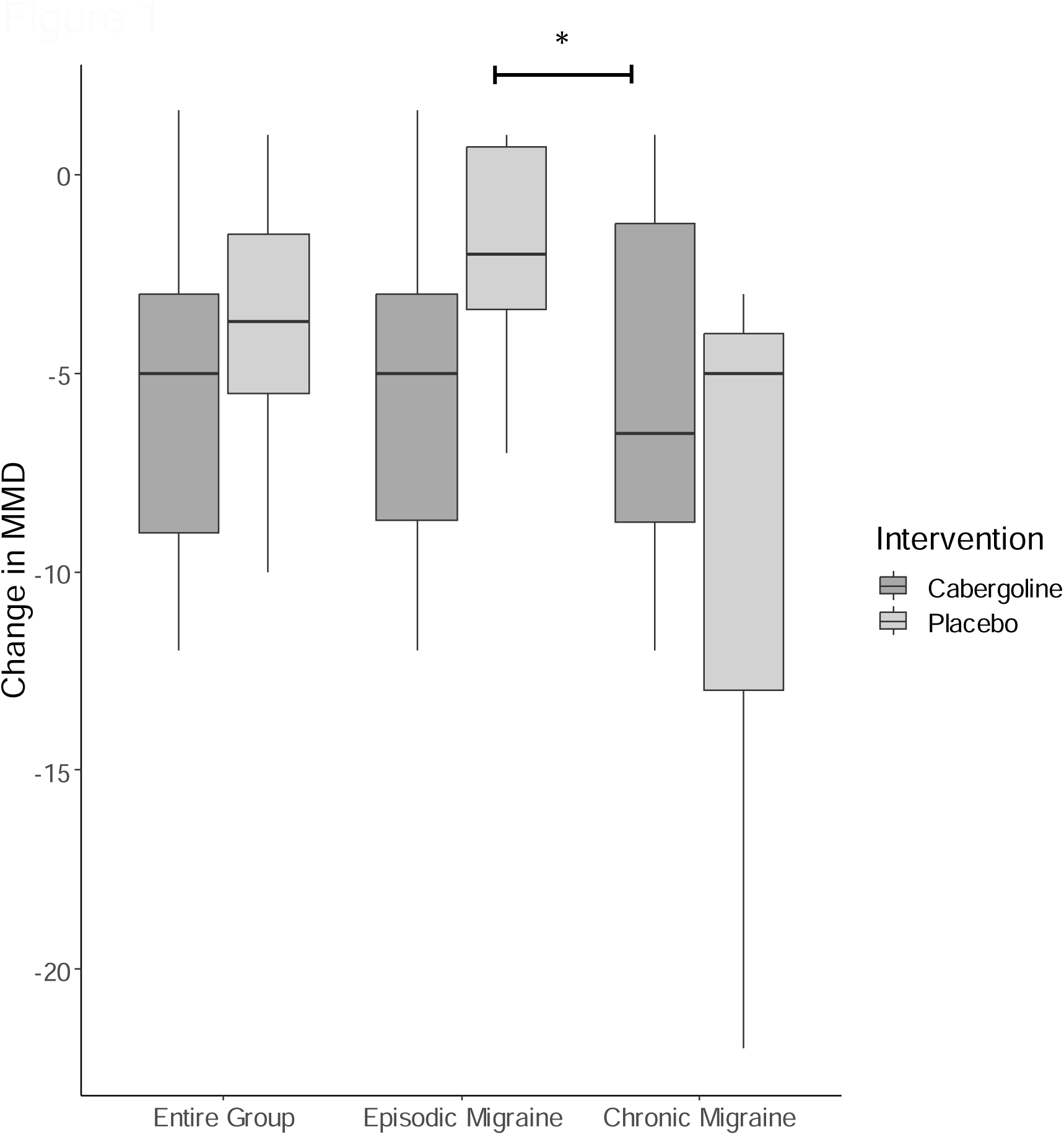
Change in number of monthly migraine days (MMD) from baseline to the final 28 days of the treatment period in the entire group of participants, in participants with episodic migraine and in participants with chronic migraine. Medians, percentiles, minimums, and maximums are presented. * p < 0.05

Among participants with episodic migraine, the odds ratio of having a day with migraine after cabergoline treatment as compared to placebo was 0.79 (95% CI: 0.65-0.95, p=0.014), whereas participants with chronic migraine had an OR of 1.3 (95% CI: 0.87-1.91, p=0.2).

### Fifty percent reduction in monthly migraine days

A 50% or greater reduction in MMD was achieved for 8 out of 17 participants in the cabergoline group, as compared with 4 out of 16 in the placebo group (p=0.28) (Figure 2). In participants with episodic migraine, 6 out of 11 participants in the cabergoline group and 1 out of 9 participants in the placebo groups obtained a reduction of at least 50% (Figure 2) (p=0.07). In participants with chronic migraine, 2 out of 6 participants in the cabergoline group and 3 out of 7 participants in the placebo group had a reduction of 50% or greater (Figure 2).

**Figure 2.**
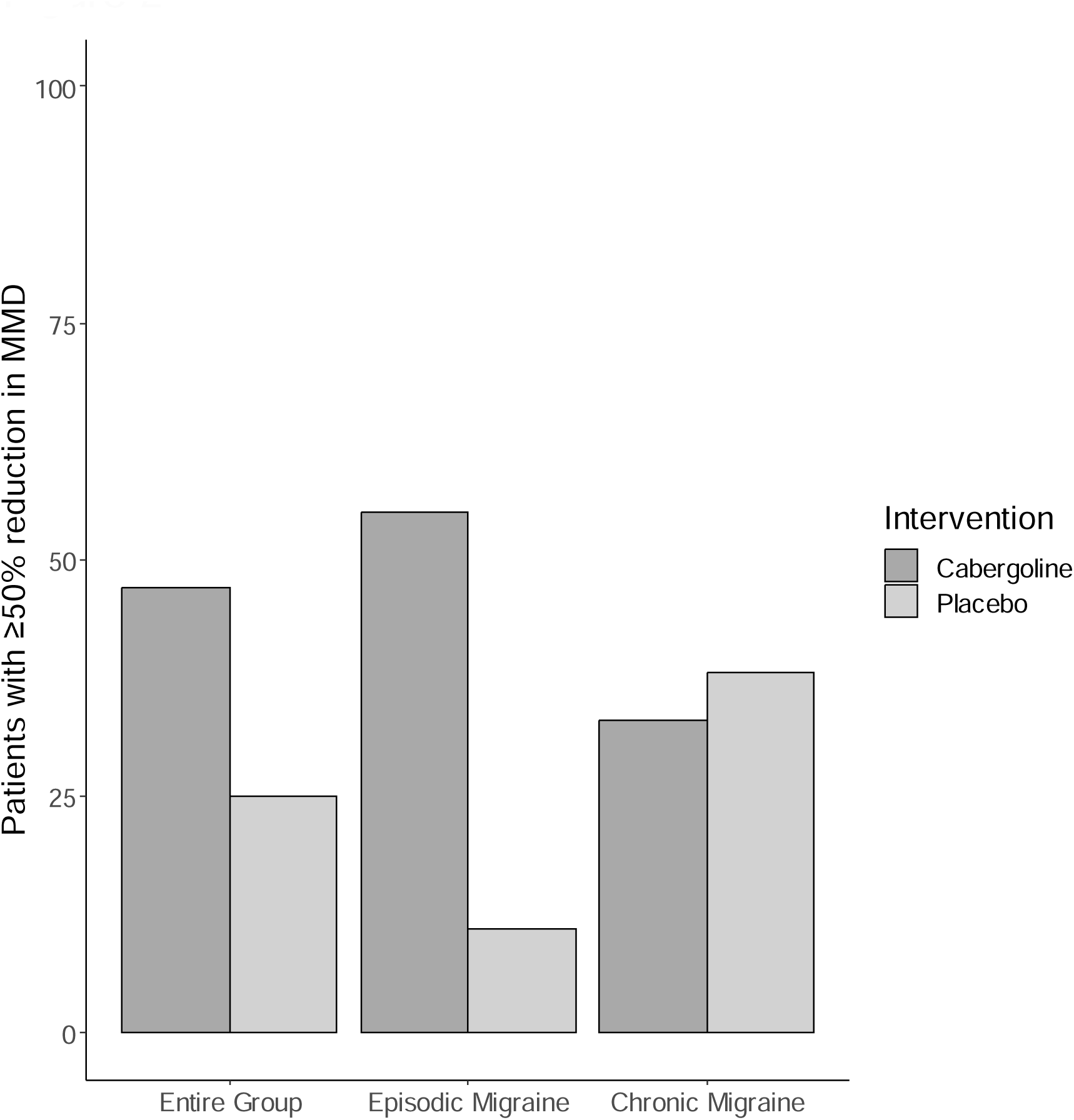
Percentage of participants with 50% reduction in monthly migraine days (MMD) in the entire group of participants, in participants with episodic migraine and in participants with chronic migraine.

### Patient Global Impression of Change (PGIC)

The PGIC score significantly increased after cabergoline compared to placebo in the entire group [median (IQR): 8 (3) vs. 5 (1.25), p=0.006], and in participants with episodic migraine [median (IQR): 8 (2.5) vs. 5 (0), p=0.01]. No significant difference was found in participants with chronic migraine [median (IQR): 7.5 (2.5) vs. 5.5 (2), p=0.2] (Figure 3). Four out of 17 participants in the cabergoline group and 11 out of 16 participants in the placebo group reported *no change* or *change for the worse* after treatment (p=0.01). The PGIC score correlated inversely with the change in MMD in all participants (r = -0.67, p<0.0001), and in participants with episodic migraine (r = -0.71, p=0.0005) and chronic migraine (r = -0.68, p=0.01).

**Figure 3.**
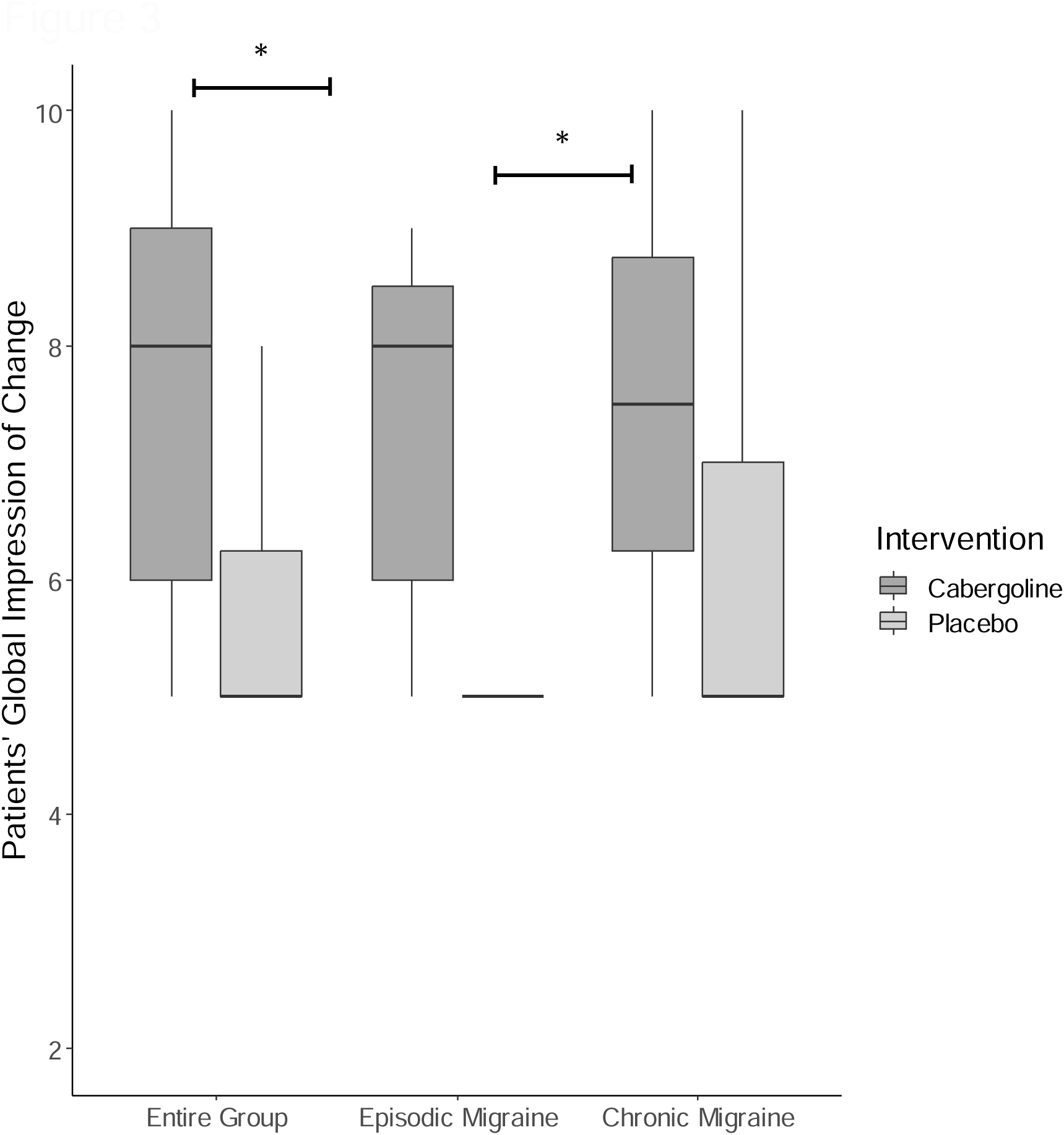
Patients’ Global Impression of Change (PGIC) in the entire group of participants, in participants with episodic migraine and in participants with chronic migraine. Medians, percentiles, minimums, and maximums are presented. * p < 0.05

### Change from baseline in MIDAS and HIT-6 score and use of acute migraine–specific medication (triptans)

No differences in use of triptans or change in either MIDAS or HIT-6 score were found between groups (Table 2).

### Participants’ best guess about treatment allocation

Twelve out of 16 participants in the cabergoline group guessed they had received active treatment as compared to 3 out of 15 in the placebo group (p=0.004). In participants with episodic migraine, 8 out of 11 in the cabergoline group guessed they received active treatment as compared to 1 out of 9 in the placebo group (p=0.01). In participants with chronic migraine, 4 out of 6 in the cabergoline group and 2 out of 7 in the placebo group perceived they had received the active treatment (p=0.2).

### Serum prolactin measurements

All participants had serum prolactin levels within the normal range and prolactin levels declined significantly after cabergoline treatment regardless of migraine phenotype (Table 2). Participants with chronic migraine had higher prolactin levels (mU/l) at baseline as compared to participants with episodic migraine [median (IQR): 204 (90.8) vs 276 (143), p=0.02], whereas there was no significant difference at follow up [median (IQR): 158 (118) vs 181 (199), p=0.2]. Prolactin levels tended to correlate with the MMD at baseline (r = 0.3, p= 0.09). No correlation between change in prolactin and change in MMD was found in participants with either episodic or chronic migraine.

### Adverse events

Seven participants in the cabergoline group and 4 participants in the placebo group experienced adverse effects (p=0.05) (Supplemental Table 1). The most common (≥ 2 participants) adverse events in the cabergoline group included headache the first day following medication, fatigue, dizziness, and obstipation, whereas headache the day following medication was the most frequent adverse event in the placebo group. In general, participants reported that the adverse events subsided during the treatment period. No serious adverse events were reported.

## Discussion

In this pilot study, cabergoline treatment in a weekly dose of 0.5 mg significantly reduced the number of MMD in episodic migraine and led to a significant global patient perceived improvement. To our knowledge, this is the first RCT to record a beneficial effect of preventive dopamine agonist treatment in migraine, which substantiates observations made in uncontrolled studies^8,9,11,14^.

Among participants with chronic migraine, a positive effect of cabergoline was less obvious, which may relate to inadequate study power or a true lack of treatment efficacy in these participants. Our study population comprised only 13 participants with chronic migraine, which is particularly noteworthy since this group commonly exhibits a lower treatment response^20^ also to CGRP-targeting therapies^21^. Migraine trials are characterized by a substantial placebo effect^22^ and the larger placebo response in participants with chronic migraine in our study may accordingly be explained by a combination of more severe symptoms and a higher expectation of pain relief^23^.

When comparing the outcome of this study to contemporary migraine trials, the efficacy of cabergoline in episodic migraine in terms of a MMD reduction of 3.6 days is superior to approved preventive treatments including CGRP-targeted therapy^24^. According to a recent review, the average reduction in MMD for anti-CGRP treatment versus placebo in episodic and chronic migraine, were 1.9 and 2.2 days respectively, whereas MMD reduction by other preventive treatments, including propranolol, metoprolol, onabotulinumtoxinA, topiramate, valproate, candesartan and amitriptyline, ranged from 0.9 to 1.7 days in episodic migraine and 1.8 to 2 days in chronic migraine^24^. Thus, the cabergoline effect observed in episodic migraine is promising but it needs to be replicated in a larger study.

Next to lack of efficacy, adverse events are the most common reason for discontinuation of preventive migraine treatment^24^. Side effects of cabergoline were mild and included transient headache, dizziness, fatigue, and obstipation. While long-term, high-dose cabergoline treatment has been associated with an increased risk of valvular heart disease in patients with Parkinson’s disease based on retrospective registry data, this has not occurred after cabergoline treatment of hyperprolactinemia using doses comparable to that of the current study^25^. In the present study, no serious adverse events were reported.

Dopamine has been implicated in the pathophysiology of migraine but its exact role remains elusive^5^. On one hand, dopaminergic symptoms, such as yawning, nausea, and gastrokinetic disturbances occur in the premonitory phase of migraine, and D_2_-like receptor antagonists may provide relief of these symptoms^5,26^. On the other hand, preclinical studies in rats indicate that dopamine attenuates nociceptive signaling in the trigeminocervical complex, which expresses dopamine receptors and plays a central role in migraine pathophysiology^27^. In line with this, clinical data suggest that dopamine agonists may exert beneficial effect in the headache phase of migraine^5–7^. Of particular interest, continuous bromocriptine treatment for one year reduced migraine frequency by 72 % in 18 of 24 women with menstrual migraine^6^. The study was, however, uncontrolled and provided limited insight into underlying mechanisms^6^.

As mentioned, dopamine agonists including bromocriptine and cabergoline are used to treat hyperprolactinemia, and it has been suggested that prolactin *per se* plays a role in migraine pathophysiology. Excursions in serum prolactin are hypothesized to trigger menstrual migraine^6^ and preclinical data in rodent models demonstrate prolactin receptor signaling to amplify migraine-like behavior^28^. Moreover, prolactin gene deletion has been shown to attenuate the CGRP pathway^28^. On the other hand, it is also argued that elevated prolactin levels observed in migraine patients is an epiphenomenon of reduced dopamine activity, and headache relief after dopamine agonist treatment in patients with hyperprolactinemia does not correlate with prolactin lowering^8^. In the present study, all participants had serum prolactin levels within the normal range at baseline, although higher levels were found in chronic migraine. Cabergoline significantly decreased prolactin levels in all participants, indicating compliance, but this was not correlated to a reduction in MMD.

The major strength of our study was the randomized, placebo-controlled design but the small sample size is a limitation, and a larger study is therefore needed to substantiate our findings. The proportion of participants correctly guessing their treatment allocation was larger in the cabergoline group. We found no evidence of untoward unblinding and the participants’ guess did not seem driven by adverse effects. The placebo effect in our study is low compared to other preventive migraine trials^22^, which speaks against increased expectation bias.

There is still an unmet need for more effective and well-tolerated preventive treatments of patients with episodic and chronic migraine. At the same time, the cost of migraine pharmacotherapies has increased dramatically, mainly due to the increased use of CGRP-targeted compounds^29^. In this context, repurposing of cabergoline would be an affordable option.

In conclusion, this study indicates that cabergoline may be effective in the preventive treatment of migraine without major adverse effects. These results merit to be tested in a larger randomized controlled trial.

## Data Availability

All data produced in the present study are available upon reasonable request to the authors

## Acknowledgement

We would like to thank Pia Buchtrup Hornbek and Lenette Egelund Pedersen for their lab assistance, and the study participants for their valuable contributions.

## References

1. Ashina M. Migraine. N Engl J Med 2020; 383(19): 1866–76.

2. Steiner TJ, Stovner LJ, Jensen R, Uluduz D, Katsarava Z, Lifting The Burden: the Global Campaign against H. Migraine remains second among the world’s causes of disability, and first among young women: findings from GBD2019. J Headache Pain 2020; 21(1): 137.

3. Barbanti P, Egeo G, Aurilia C, et al. Predictors of response to anti-CGRP monoclonal antibodies: a 24-week, multicenter, prospective study on 864 migraine patients. J Headache Pain 2022; 23(1): 138.

4. Kelley NE, Tepper DE. Rescue therapy for acute migraine, part 1: triptans, dihydroergotamine, and magnesium. Headache 2012; 52(1): 114–28.

5. Charbit AR, Akerman S, Goadsby PJ. Dopamine: what’s new in migraine? Curr Opin Neurol 2010; 23(3): 275–81.

6. Herzog AG. Continuous bromocriptine therapy in menstrual migraine. Neurology 1997; 48(1): 101–2.

7. Mascia A, Afra J, Schoenen J. Dopamine and migraine: a review of pharmacological, biochemical, neurophysiological, and therapeutic data. Cephalalgia : an international journal of headache 1998; 18(4): 174–82.

8. Kallestrup MM, Kasch H, Osterby T, Nielsen E, Jensen TS, Jorgensen JO. Prolactinoma-associated headache and dopamine agonist treatment. Cephalalgia 2014; 34(7): 493–502.

9. Bussone G, Usai S, Moschiano F. How to investigate and treat: headache and hyperprolactinemia. Curr Pain Headache Rep 2012; 16(4): 365–70.

10. Kreitschmann-Andermahr I, Siegel S, Weber Carneiro R, Maubach JM, Harbeck B, Brabant G. Headache and pituitary disease: a systematic review. Clin Endocrinol (Oxf) 2013; 79(6): 760–9.

11. Bosco D, Belfiore A, Fava A, et al. Relationship between high prolactin levels and migraine attacks in patients with microprolactinoma. J Headache Pain 2008; 9(2): 103–7.

12. Al-Karagholi MA-M, Kalatharan V, Ghanizada H, Gram C, Dussor G, Ashina M. Prolactin in headache and migraine: A systematic review of clinical studies. Cephalalgia 2023; 43(2): 03331024221136286.

13. Noori-Zadeh A, Karamkhani M, Seidkhani-Nahal A, Khosravi A, Darabi S. Evidence for hyperprolactinemia in migraineurs: a systematic review and meta-analysis. Neurol Sci 2020; 41(1): 91–9.

14. Cavestro C, Rosatello A, Marino MP, Micca G, Asteggiano G. High prolactin levels as a worsening factor for migraine. J Headache Pain 2006; 7(2): 83–9.

15. Melmed S, Casanueva FF, Hoffman AR, et al. Diagnosis and treatment of hyperprolactinemia: an Endocrine Society clinical practice guideline. The Journal of clinical endocrinology and metabolism 2011; 96(2): 273–88.

16. Kvernmo T, Härtter S, Burger E. A review of the receptor-binding and pharmacokinetic properties of dopamine agonists. Clinical therapeutics 2006; 28(8): 1065–78.

17. Harris PA, Taylor R, Minor BL, et al. The REDCap consortium: Building an international community of software platform partners. J Biomed Inform 2019; 95: 103208.

18. Headache Classification Committee of the International Headache Society (IHS) The International Classification of Headache Disorders, 3rd edition. Cephalalgia 2018; 38(1): 1–211.

19. Carvalho GF, Luedtke K, Braun T. Minimal important change and responsiveness of the Migraine Disability Assessment Score (MIDAS) questionnaire. J Headache Pain 2021; 22(1): 126.

20. Mungoven TJ, Henderson LA, Meylakh N. Chronic Migraine Pathophysiology and Treatment: A Review of Current Perspectives. Front Pain Res (Lausanne) 2021; 2: 705276.

21. Raffaelli B, Fitzek M, Overeem LH, Storch E, Terhart M, Reuter U. Clinical evaluation of super-responders vs. non-responders to CGRP(-receptor) monoclonal antibodies: a real-world experience. J Headache Pain 2023; 24(1): 16.

22. Evans K, Romero H, Spierings EL, Katz N. The relation between the placebo response, observed treatment effect, and failure to meet primary endpoint: A systematic review of clinical trials of preventative pharmacological migraine treatments. Cephalalgia 2021; 41(2): 247–55.

23. Sanders AE, Slade GD, Fillingim RB, Ohrbach R, Arbes SJ, Jr., Tchivileva IE. Effect of Treatment Expectation on Placebo Response and Analgesic Efficacy: A Secondary Aim in a Randomized Clinical Trial. JAMA Netw Open 2020; 3(4): e202907.

24. Vandervorst F, Van Deun L, Van Dycke A, et al. CGRP monoclonal antibodies in migraine: an efficacy and tolerability comparison with standard prophylactic drugs. J Headache Pain 2021; 22(1): 128.

25. Steeds R, Stiles C, Sharma V, Chambers J, Lloyd G, Drake W. Echocardiography and monitoring patients receiving dopamine agonist therapy for hyperprolactinaemia: A joint position statement of the British Society of Echocardiography, the British Heart Valve Society and the Society for Endocrinology. Clin Endocrinol (Oxf) 2019; 90(5): 662–9.

26. Barbanti P, Aurilia C, Egeo G, Fofi L, Guadagni F, Ferroni P. Dopaminergic symptoms in migraine: A cross-sectional study on 1148 consecutive headache center-based patients. Cephalalgia 2020; 40(11): 1168–76.

27. Bergerot A, Storer RJ, Goadsby PJ. Dopamine inhibits trigeminovascular transmission in the rat. Ann Neurol 2007; 61(3): 251–62.

28. Al-Karagholi MA, Kalatharan V, Ghanizada H, Dussor G, Ashina M. Prolactin in headache and migraine: A systematic review of preclinical studies. Headache 2023; 63(5): 577–84.

29. Nguyen JL, Munshi K, Peasah SK, et al. Trends in utilization and costs of migraine medications, 2017-2020. J Headache Pain 2022; 23(1): 111.

