## Supplementary figures and images for "Cabergoline as a Preventive Migraine Treatment: An Investigator-Initiated Randomized Controlled Trial"

### Supplemental Figure 1

**Supplementary Figure 1.**

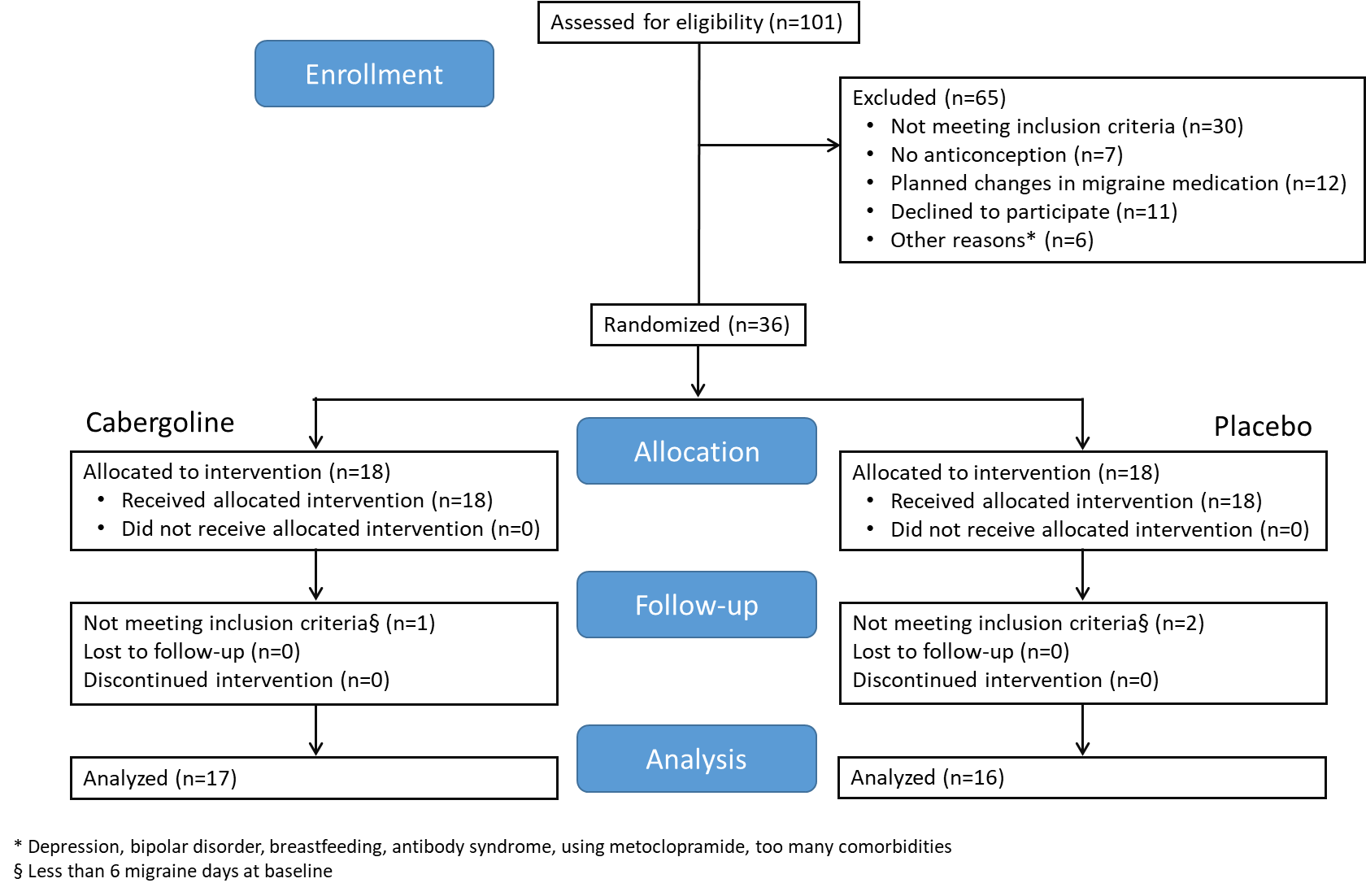
