## Supplemental Table 1 for "Cabergoline as a Preventive Migraine Treatment: An Investigator-Initiated Randomized Controlled Trial"

**Supplementary Table 1. Adverse Events Reported during the Treatment Phase.**

|  | Cabergoline (n=18) | Placebo  (n=18) |
| --- | --- | --- |
| Adverse event – no. | 7 | 4 |
| Headache the day following medication | 4 | 2 |
| Fatigue | 4 | - |
| Dizziness | 2 | - |
| Obstipation | 2 | - |
| Sweating | 1 | - |
| Stabbing chest pain | 1 | - |
| Weight gain | 1 | - |
| Headache | - | 1 |
| Loss of appetite | - | 1 |

More than one adverse event could be reported by a participant.
